# Accuracy and Consistency of Frontier LLMs on Orthodontic Diagnostic Tasks: A Repeated-Trial Comparison

**DOI:** 10.64898/2026.05.17.26353409

**Authors:** Kang Wan Jing, Jonathan Sim, Eugene Loh Eu-Min, Arthur Lim Chong Yang, Kelvin Weng Chiong Foong

**Affiliations:** Faculty of Dentistry, National University of Singapore, Singapore; Department of Philosophy, National University of Singapore, Singapore; The Implant and Oral Surgery Centre, 10 Sinaran Drive #11-25/26, Singapore 307506

**Author notes:** **Corresponding author:** Kelvin Weng Chiong Foong, Faculty of Dentistry, National University of Singapore, 9, Lower Kent Ridge Road, Singapore.

**Keywords:** Large Language Models, Orthodontics, Cephalometry, Reproducibility of Results, Decision Support Systems, Clinical, Sensitivity and Specificity

## Abstract

**Importance:** Large language models are increasingly explored as clinical decision-support tools in orthodontics, yet existing evaluations have been confined to knowledge-based question answering where reported accuracy ranges from 18% to 100%. No study has evaluated performance on the computational and classificatory tasks that define daily diagnostic work. Furthermore, 84.3% of published healthcare large language model studies fail to report the number of repeated queries performed, leaving output stochasticity unexamined.

**Objective:** To compare the diagnostic accuracy and output consistency of three frontier reasoning-enhanced large language models, namely, ChatGPT 5.4 (Thinking), Gemini 3 (Thinking), and Claude Opus 4.6 (Extended Thinking), on Bolton analysis, Index of Orthodontic Treatment Need-Dental Health Component (IOTN-DHC) classification, space analysis, and lateral cephalometric interpretation.

**Methods:** In this comparative cross-sectional study with a repeated-measures design, each model, accessed through its respective consumer-facing web interfaces under default provider settings rather than through application programming interfaces, processed 200 purpose-built items (50 per task) across four independent trials, yielding 2,400 observations. Responses were scored against a pre-established reference standard by two independent raters using strict binary exact-match criteria. Accuracy was reported with exact binomial 95% confidence intervals. Inter-model comparisons used Cochran’s Q test with post-hoc McNemar’s tests and Bonferroni correction. A supplementary context-rich prompting evaluation was conducted on 40 items (480 observations).

**Results:** Claude Opus 4.6 (Extended Thinking) achieved the highest accuracy (99.0%; 95% CI: 96.4–99.9%), followed by Gemini 3 (Thinking) (95.5%; 91.6–98.1%) and ChatGPT 5.4 (Thinking) (94.0%; 89.8–96.9%) (Cochran’s Q = 6.87, p = 0.032). Each model exhibited distinct, non-overlapping error profiles concentrated at the normal–abnormal classification boundary. An accuracy–consistency paradox emerged: the most accurate model was the least consistent (93.0%), while the least accurate was the second-most consistent (98.0%). Context-rich prompting eliminated all errors across all three models.

**Interpretation:** Frontier reasoning large language models achieved high overall accuracy on orthodontic diagnostic tasks but retained concealed, task-specific vulnerabilities detectable only through repeated-trial evaluation. An accuracy-consistency paradox, in which the most accurate model was the least consistent, demonstrates that single-trial evaluations cannot characterise clinical risk. The reasoning modes were associated with high arithmetic accuracy but did not compensate for imprecise parametric knowledge on classification tasks; however, the absence of a non-thinking baseline means this association cannot be attributed to the thinking mode itself. Context-rich prompting eliminated all errors on synthetic data but should be regarded as a necessary yet insufficient prerequisite for clinical deployment pending prospective validation on real patient data.

## Introduction

Artificial intelligence has an extensive trajectory within orthodontic research, progressing from rule-based expert systems to deep learning architectures capable of automating cephalometric landmark identification and treatment need assessment.^1^ The most extensively studied applications have involved image-based analyses, where supervised systems achieve clinically meaningful accuracy when tasks can be framed as structured pattern recognition problems with well-defined ground truth.^2^ The emergence of large language models (LLMs) represents a fundamentally different paradigm, in which general-purpose systems trained on text that can, in principle, address any language-mediated clinical task without task-specific training.

A systematic review of 519 healthcare LLM studies found that accuracy was the predominant evaluation dimension, yet only 5% used real patient data and critical dimensions including consistency and reproducibility remained inadequately assessed.^3^ In orthodontics specifically, existing evaluations have been confined to informational question-answering, where reported accuracy ranges widely from 18% to 100% depending on the model, task, and evaluation method.^4,5,6^ No published study has systematically evaluated LLMs on the core computational and classificatory tasks that define daily orthodontic diagnostic work: Bolton analysis, Index of Orthodontic Treatment Need–Dental Health Component (IOTN-DHC) classification, space analysis, and lateral cephalometric interpretation.

This gap is significant because these tasks are precisely those most likely to be delegated to AI tools in clinical practice, and the risk of automation bias is compounded by theoretical evidence that hallucinations in large language models are structurally inevitable.^7^ They appear simple, they are time-consuming, and they involve well-defined numerical procedures that seem amenable to automation. Yet they require precise arithmetic, accurate application of thresholds, and integrative clinical reasoning, each of which lies within a known area of LLM vulnerability.^8,9^ Furthermore, a striking 84.3% of published LLM studies in leading medical journals failed to disclose the number of repeated queries performed,^10^ representing a critical reproducibility gap, given that output stochasticity has been documented across multiple clinical specialities and persists even under maximally deterministic settings.^11,12^

Reasoning-enhanced LLMs, which engage in extended internal deliberation before producing a response, have shown remarkable promise on mathematical benchmarks^13,14^ and clinical evaluation in emergency medicine,^15^ but the application of this paradigm to orthodontic diagnostic computation has not been studied. The distinction between parametric knowledge (information encoded in model weights during pre-training) and in-context knowledge (information provided within the prompt) is central to interpreting LLM behaviour on threshold-dependent clinical tasks,^16^ because prompt design may be as important as model selection in determining output accuracy and reliability.^17,18^

This study provides the first reproducibility-aware, head-to-head comparison of three frontier reasoning-enhanced LLMs, namely, ChatGPT 5.4 (Thinking), Gemini 3 (Thinking), and Claude Opus 4.6 (Extended Thinking), on Bolton analysis, IOTN-DHC classification, space analysis, and lateral cephalometric interpretation. We address three research questions: (1) How accurate are these models on rule-based classification and numeric computation tasks in orthodontic diagnosis? (2) How consistent are their outputs across repeated independent trials? (3) Do the models differ significantly in accuracy and consistency, and which task categories remain error-prone?

## Methodology

### Study design

This comparative cross-sectional study evaluated the accuracy and output consistency of ChatGPT 5.4 (Thinking) (OpenAI), Gemini 3 (Thinking) (Google DeepMind), and Claude Opus 4.6 (Extended Thinking) (Anthropic) on four orthodontic diagnostic tasks. Each model processed every test item across four independent trials in a repeated-measures design, yielding 2,400 observations (3 models × 4 tasks × 50 items × 4 trials). As the investigation involved only computational evaluation of commercially available AI systems using synthetic clinical data containing no identifiable personal health information, institutional review board approval was not required.

### Test item construction and reference standard

A dataset of 200 items was constructed, comprising 50 items for each of four task categories: Bolton analysis, IOTN-DHC classification,^19^ space analysis, and lateral cephalometric interpretation. All items were purpose-built using synthetic numerical data spanning a wide clinically relevant range. An a priori sample size calculation determined that a minimum of 45 items per task would be required to detect a significant difference at 90% power and 5% significance after Bonferroni adjustment; 50 items per task were therefore constructed.

Bolton analysis tested multi-step decimal arithmetic and ratio computation against Bolton’s normative values (anterior ratio 77.2 ± 1.65%; overall ratio 91.3 ± 1.91%).^20,21^ IOTN-DHC classification assessed hierarchical rule-based classification using the five-grade system of Brook and Shaw.^19^ Space analysis evaluated numerical computation with sign-dependent reasoning based on the tooth size-arch length discrepancy concept.^22^ Lateral cephalometric interpretation tested integrative synthesis of multiple numerical parameters against the southern Chinese cephalometric standards of Cooke and Wei,^23^ incorporating SNA, SNB, ANB, Wits appraisal,^24,25^ vertical dimension indices, and incisor inclination measurements.

Reference standard answers for all 200 items were independently calculated and verified by two investigators using manual computation with a standard scientific calculator, cross-checked against the published normative values and classification criteria.

### Model selection and configuration

The three models were selected to represent each major commercially available model family, each with an advanced reasoning mode, accessed through their respective consumer-facing web interfaces (chat.openai.com, gemini.google.com, claude.ai) rather than through application programming interfaces. This reflected the access modality most likely to be used by practising orthodontists. No modifications were made to default model parameters. The “Thinking” or “Extended Thinking” mode was activated for each model prior to every session. All primary data were collected on 7 March 2026.

### Prompt design and data collection

A single standardised prompt template was developed for each task category, iteratively refined through pilot testing. Each template presented the numerical data in structured format, specified the required output, and identified the normative reference values. For each trial, a new chat session was initiated to ensure independence. Model outputs were recorded verbatim into a structured data collection spreadsheet.

### Supplementary evaluation: context-rich prompting

The error patterns identified during the primary evaluation raised the question of whether the observed failures reflected fundamental reasoning limitations or imprecise parametric knowledge amenable to correction. A post-hoc supplementary evaluation re-administered 40 items and yielded 480 observations (10 items per task x 4 tasks x 3 models x 4 trials) under a context-rich prompting condition that embedded explicit classification rules, numerical boundaries, and decision hierarchies within the prompt text. All other elements remained identical to the primary condition. Supplementary data were collected on 18 March 2026.

### Outcome scoring

Each output was scored as correct (1) or incorrect (0) against the reference standard. For Bolton analysis, both ratios were required to fall within ±0.5 percentage points of the reference values with correct discrepancy identification. For IOTN-DHC, the exact grade and sub-classification code were required. For space analysis, the discrepancy for both arches was required with correct magnitude and sign. For lateral cephalometric interpretation, the integrated summary classification was required to match the reference standard. Scoring was performed by two independent raters, blinded to each other’s scores. Intra-rater reliability was assessed by repeating the complete scoring after four days.

### Statistical analysis

Accuracy was reported as a percentage with exact binomial 95% confidence intervals using modal scores. Cochran’s Q test compared the three models on matched items, with post-hoc McNemar’s tests and Bonferroni correction (α = 0.0167) against a pre-specified minimum clinically important difference (MCID) of 10 percentage points. Response consistency was assessed as the proportion of items with perfect four-trial agreement; within-model Cochran’s Q tests assessed trial-to-trial variation. Inter-model agreement was quantified using Cohen’s kappa, Fleiss’ kappa, and three-way agreement proportions. A generalised estimating equations (GEE) model with binomial family, logit link, and exchangeable within-item correlation was fitted to all 2,400 observations. Confusion matrices with per-class sensitivity and positive predictive value (PPV) were constructed for categorical tasks. All tests used α = 0.05 unless otherwise specified.

## Results

### Overall and task-specific accuracy

Claude Opus 4.6 (Extended Thinking) achieved the highest overall accuracy at 99.0% (198/200; 95% CI: 96.4 to 99.9%), followed by Gemini 3 (Thinking) at 95.5% (191/200; 91.6 to 98.1%) and ChatGPT 5.4 (Thinking) at 94.0% (188/200; 89.8 to 96.9%). The overall Cochran’s Q test was significant (Q = 6.87, df = 2, p = 0.032). Task-specific accuracy is presented in Table 1.

**Table 1.** Accuracy (% correct out of 50 items) by task category and trial for each model.

| Task | Trial 1 | Trial 2 | Trial 3 | Trial 4 | Modal | 95% CI |
| --- | --- | --- | --- | --- | --- | --- |
| **ChatGPT 5.4 (Thinking)** |  |  |  |  |  |  |
| Bolton Analysis | 100 | 100 | 100 | 100 | **100.0** |  |
| IOTN-DHC | 100 | 98 | 100 | 100 | **100.0** |  |
| Space Analysis | 96 | 98 | 100 | 100 | **100.0** |  |
| Lateral Cephalometric | 76 | 76 | 76 | 76 | **76.0** |  |
| Overall |  |  |  |  | **94.0** | 89.8 to 96.9 |
| **Gemini 3 (Thinking)** |  |  |  |  |  |  |
| Bolton Analysis | 100 | 100 | 100 | 100 | **100.0** |  |
| IOTN-DHC | 86 | 86 | 88 | 88 | **88.0** |  |
| Space Analysis | 98 | 98 | 98 | 98 | **98.0** |  |
| Lateral Cephalometric | 96 | 96 | 96 | 96 | **96.0** |  |
| Overall |  |  |  |  | **95.5** | 91.6 to 98.1 |
| **Claude Opus 4.6 (Extended Thinking)** |  |  |  |  |  |  |
| Bolton Analysis | 84 | 92 | 82 | 96 | **96.0** |  |
| IOTN-DHC | 92 | 94 | 100 | 100 | **100.0** |  |
| Space Analysis | 100 | 100 | 100 | 100 | **100.0** |  |
| Lateral Cephalometric | 100 | 100 | 100 | 100 | **100.0** |  |
| Overall |  |  |  |  | **99.0** | 96.4 to 99.9 |

ChatGPT 5.4 (Thinking) achieved 100% accuracy on Bolton analysis across all four trials but only 76% on lateral cephalometric interpretation, with the identical 12 items misclassified in every trial. Gemini 3 (Thinking) achieved 100% on Bolton analysis, but IOTN-DHC classification ranged from 86% to 88%, with six items systematically misclassified. Claude Opus 4.6 (Extended Thinking) achieved 100% on both space analysis and lateral cephalometric interpretation across all trials but showed the greatest trial-to-trial variability on Bolton analysis (82 to 96%).

Task-specific Cochran’s Q tests revealed significant inter-model divergence for IOTN-DHC classification (Q = 12.000, p = 0.002) and lateral cephalometric interpretation (Q = 17.714, p < 0.001), but not for Bolton analysis (p = 0.135) or space analysis (p = 0.368). Post-hoc pairwise comparison showed a significant difference only between ChatGPT 5.4 (Thinking) and Claude Opus 4.6 (Extended Thinking) (χ² = 5.786, p = 0.016, Δ = 5.0 percentage points). No pairwise difference reached the pre-specified MCID of 10 percentage points.

### Response consistency

Gemini 3 (Thinking) demonstrated the highest overall consistency at 99.5% (199/200 items perfectly consistent across four trials), followed by ChatGPT 5.4 (Thinking) at 98.0% and Claude Opus 4.6 (Extended Thinking) at 93.0% (Table 2). ChatGPT 5.4 (Thinking)’s high consistency included its systematic errors: the 12 misclassified cephalometric items were reproduced identically in every trial. Within-model Cochran’s Q tests confirmed that Claude Opus 4.6 (Extended Thinking) was the only model with statistically significant trial-to-trial variation, on Bolton analysis (Q = 10.622, p = 0.014) and IOTN-DHC classification (Q = 10.200, p = 0.017).

**Table 2.** Response agreement across all four trials, by model and task (n = 50 items per task). Values represent the number (%) of items with identical outcomes across all four trials.

| Model | Bolton | IOTN | Space | Ceph. | Overall |
| --- | --- | --- | --- | --- | --- |
| Gemini 3 (Thinking) | 50 (100) | 49 (98) | 50 (100) | 50 (100) | **99.5%** |
| ChatGPT 5.4 (Thinking) | 50 (100) | 49 (98) | 47 (94) | 50 (100) | **98.0%** |
| Claude Opus 4.6 (ET) | 40 (80) | 46 (92) | 50 (100) | 50 (100) | **93.0%** |

ET = Extended Thinking.

### Inter-model agreement and item-level error analysis

Across the entire 200-item dataset, no two models failed on the same item. Every error was model-specific: ChatGPT 5.4 (Thinking) accounted for 12 lateral cephalometric misclassifications, Gemini 3 (Thinking) for 6 IOTN-DHC sub-classification errors, and Claude Opus 4.6 (Extended Thinking) for 2 Bolton analysis items (on modal score). Three-way agreement was 88.5% overall. A consensus-based multi-model verification approach would have achieved 100% accuracy on this dataset.

### Confusion matrix analysis

ChatGPT 5.4 (Thinking)’s 12 lateral cephalometric errors reflected a unidirectional Class II overclassification bias (Table 3). Nine items with ANB values of 4-5°, lying at the upper end of the Class I range of 1-5°, were classified as “Mild Class II”, even though that category does not exist in the reference taxonomy. Three items with ANB of 0° (below the Class I lower boundary of 1°) were misclassified as Class I. Sensitivity was 40.0% for High angle Class I and 50.0% for Class III (Table 5). A phantom “Mild Class II” column contained six items with no true positives (PPV = 0%).

**Table 3.** Confusion matrix for lateral cephalometric classification by ChatGPT 5.4 (Thinking) (modal score, n = 50 items). Rows represent the reference standard class; columns represent the predicted class. Cl = Class; HA = High angle; LA = Low angle; M = Mild.

| Reference | CI I | CI II | CI III | HA-I | HA-II | HA-III | LA-I | M-II |
| --- | --- | --- | --- | --- | --- | --- | --- | --- |
| CI I (14) | 10 | 0 | 0 | 0 | 0 | 0 | 0 | 4 |
| CI II (9) | 0 | 9 | 0 | 0 | 0 | 0 | 0 | 0 |
| CI III (6) | 3 | 0 | 3 | 0 | 0 | 0 | 0 | 0 |
| HA-I (10) | 0 | 0 | 0 | 4 | 4 | 0 | 0 | 2 |
| HA-II (5) | 0 | 0 | 0 | 0 | 5 | 0 | 0 | 0 |
| HA-III (4) | 0 | 0 | 0 | 0 | 0 | 4 | 0 | 0 |
| LA-I (2) | 0 | 0 | 0 | 0 | 0 | 0 | 1 | 0 |

Gemini 3 (Thinking)’s six IOTN-DHC errors were confined within Grade 4: the model correctly assigned the severity grade but misidentified the defining sub-classification (Table 4). Sensitivity was 50.0% for sub-category 4a and 75.0% for 4d (Table 5). No errors crossed grade boundaries. Claude Opus 4.6 (Extended Thinking) achieved perfect confusion matrices for both categorical tasks (100% sensitivity and PPV across all classes).

**Table 4.**
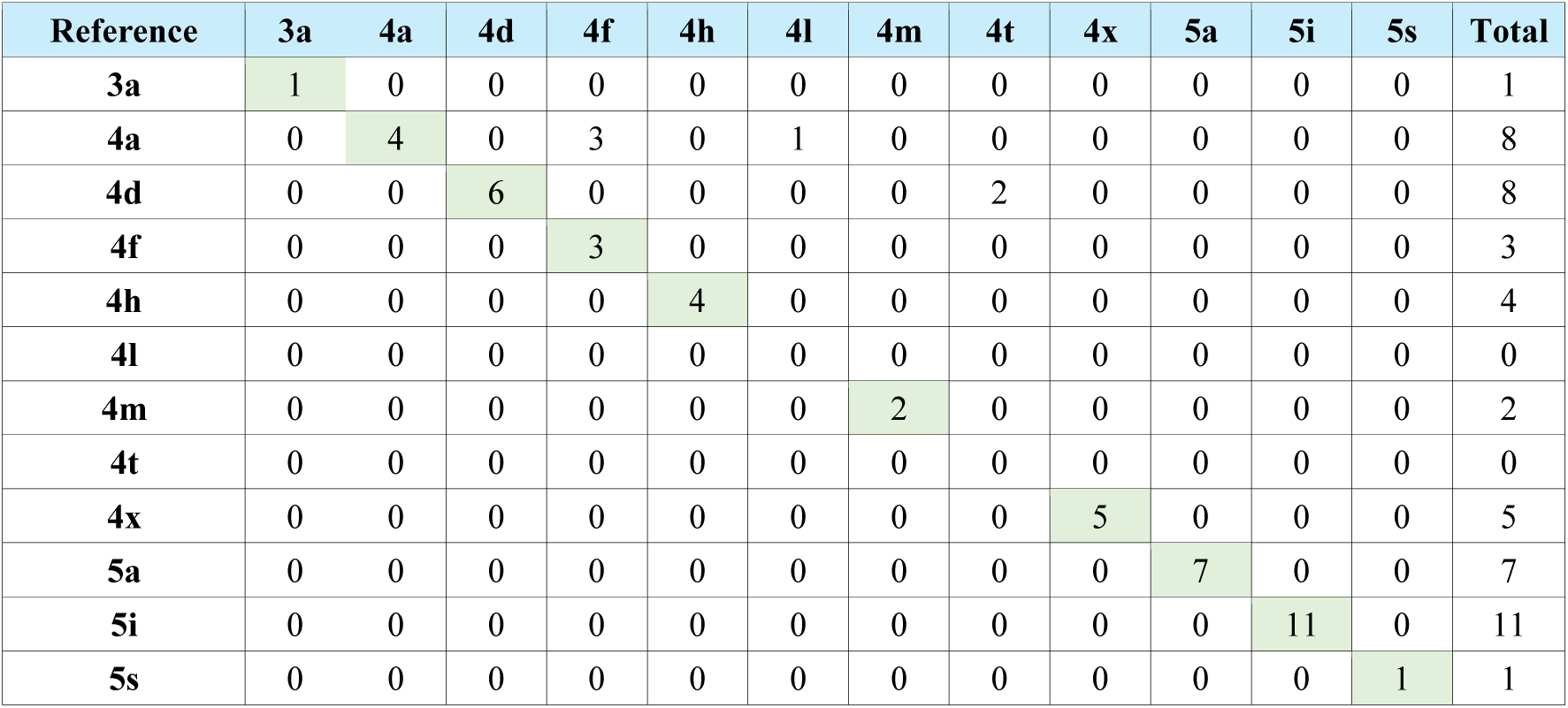
Confusion matrix for IOTN-DHC classification by Gemini 3 (Thinking) (modal score, n = 50 items). ChatGPT 5.4 (Thinking) and Claude Opus 4.6 (Extended Thinking) achieved perfect classification (50/50) and are omitted.

**Table 5.** Per-class sensitivity and positive predictive value (PPV) for lateral cephalometric (LC) and IOTN-DHC classification tasks. Only models with at least one misclassification are shown. Claude Opus 4.6 (Extended Thinking) achieved 100% for all classes on both tasks.

| Task | Model | Class | n | Sens. (%) | PPV (%) |
| --- | --- | --- | --- | --- | --- |
| LC | ChatGPT 5.4 | CI I | 14 | 71.4 | 76.9 |
| LC | ChatGPT 5.4 | CI III | 6 | 50.0 | 100.0 |
| LC | ChatGPT 5.4 | HA-I | 10 | 40.0 | 100.0 |
| LC | ChatGPT 5.4 | HA-II | 5 | 100.0 | 55.6 |
| LC | ChatGPT 5.4 | LA-I | 2 | 50.0 | 100.0 |
| LC | Gemini 3 | CI I | 14 | 92.9 | 100.0 |
| LC | Gemini 3 | CI II | 9 | 88.9 | 100.0 |
| IOTN | Gemini 3 | 4a | 8 | 50.0 | 100.0 |
| IOTN | Gemini 3 | 4d | 8 | 75.0 | 100.0 |
| IOTN | Gemini 3 | 4f | 3 | 100.0 | 50.0 |

Sens. = Sensitivity. Only classes with imperfect performance are shown; all unlisted classes achieved 100% for both metrics.

### Generalised estimating equations and reliability

The GEE model did not detect significant inter-model effects (Gemini OR = 1.39, 95% CI: 0.59–3.29, p = 0.448; Claude OR = 1.78, 0.81–3.92, p = 0.150). A model with model-by-task interaction could not be fitted due to complete separation in four cells with 100% accuracy. Intra-rater reliability was excellent (κ ≥ 0.918); inter-rater agreement was ≥99.0%.

### Context-rich prompting

Under context-rich prompting, all three models achieved 100% accuracy and 100% consistency across all four task categories (480 observations). ChatGPT 5.4 (Thinking)’s deterministic cephalometric misclassifications, Gemini 3 (Thinking)’s IOTN-DHC sub-classification errors, and Claude Opus 4.6 (Extended Thinking)’s stochastic Bolton variability were all eliminated.

## Discussion

This study provides the first systematic, reproducibility-aware evaluation of frontier reasoning LLMs on core orthodontic diagnostic computations. All three models achieved high overall accuracy (94.0 to 99.0%), but beneath these headline figures lies a complex pattern of task-specific vulnerabilities, divergent error profiles, and a previously undocumented trade-off between accuracy and consistency.

The overall accuracy levels substantially exceed those reported in previous orthodontic LLM evaluations, where ChatGPT-4 achieved entirely correct responses in only 40.5% of interceptive orthodontics cases^4^ and crowding classification from intraoral images reached only 50%.^6^ The markedly superior performance likely reflects two factors: the use of the latest reasoning-enhanced models, and the text-based, numerically specified task format that removed the confound of visual interpretation known to degrade LLM performance by approximately 10 percentage points across model families.^26^

The most striking finding was ChatGPT 5.4 (Thinking)’s systematic failure on lateral cephalometric interpretation (76% accuracy), with 12 items misclassified with perfect determinism across all four trials. Qualitative error analysis revealed a dual ANB threshold error: nine items with ANB values of 4-5° were over-classified as “Mild Class II” because the model applied a threshold at the ANB mean (3°) rather than at the upper limit of the normal range (5°), whilst three items with ANB 0° were under-classified as Class I. The thinking mode allocated additional inference-time computation to every item, yet this deliberation did not correct the underlying threshold error; the model reasoned carefully and consistently toward the wrong answer, a pattern that might be termed rigorous incorrectness. The term “Mild Class II” does not appear in formal cephalometric classification systems^23,25^ but is widely used in informal clinical discourse across the internet, which forms part of the unstructured text on which LLMs are trained. The model appears to have internalised this label without acquiring the corresponding rule that ANB values within one standard deviation of the mean are classified as Class I. This represents a form of hallucination^27^ in which the output is linguistically plausible but factually unsupported by the classificatory framework.

Gemini 3 (Thinking)’s six IOTN-DHC errors reveal a qualitatively different failure mode. All six items were correctly assigned to Grade 4, but the sub-classification letter was incorrect, reflecting failure in applying the worst-feature hierarchy when multiple Grade 4 features coexisted. Whilst these errors would not alter treatment eligibility, they indicate a deficiency in the granularity of clinical rule application that would compromise referral documentation.

Claude Opus 4.6 (Extended Thinking)’s Bolton errors demonstrated a third failure mode: stochastic arithmetic variability, with accuracy ranging from 82% to 96% across trials on items involving minor decimal discrepancies of 0.5 percentage points. This pattern is consistent with the non-deterministic token-sampling process inherent to autoregressive architectures and suggests that extended thinking, by generating longer deliberation traces with more sampled tokens, may paradoxically increase arithmetic noise as a byproduct of deeper reasoning.^13^

Perhaps the most novel contribution is the documentation of an accuracy-consistency paradox. The most accurate model (Claude, 99.0%) demonstrated the lowest consistency (93.0%), whilst the least accurate (ChatGPT, 94.0%) exhibited the second-highest consistency (98.0%) because its errors were perfectly deterministic. This paradox carries direct implications for patient safety: a single-trial evaluation cannot distinguish a model that is reliably correct from one that is reliably wrong. The overwhelming majority of published LLM evaluations are structurally unable to detect this pattern.^10^

Across all four tasks, errors were concentrated at or near the normal–abnormal classification boundary, forming a gradient from cross-boundary misclassification in cephalometric interpretation, through within-grade discrimination errors in IOTN-DHC, to sub-threshold arithmetic noise in Bolton analysis. This suggests that LLMs are most vulnerable in tasks requiring precise threshold-based judgements and least vulnerable in straightforward arithmetic with unambiguous clinical direction.

The clinical significance of these findings cannot be separated from the problem of automation bias.^7^ ChatGPT 5.4 (Thinking) achieved 100% accuracy on Bolton analysis, yet the same model, with identical confidence and zero uncertainty signalling, systematically misclassified 24% of cephalometric items, with 9 of 50 items (18%) receiving an incorrect sagittal classification capable of altering the treatment plan. A clinician who has verified the model’s flawless computational performance will rationally develop trust in its diagnostic capabilities, and this earned trust provides no basis for suspecting that the same model’s cephalometric interpretation is systematically wrong. This asymmetry between perceived and actual reliability across task domains constitutes the primary clinical risk identified by this study.

Two remediation strategies emerged from the findings. One was multi-model verification. Across the entire dataset, no single item was misclassified by more than one model, suggesting that consensus-based querying would have identified and corrected every error. The other was context-rich prompting, which yielded 100% accuracy and consistency across all three models by embedding explicit classification rules within the prompt. The elimination of all errors indicates that the original failures reflected imprecise parametric knowledge rather than a fundamental limitation in reasoning.^16,17,18^ These errors were corrected once authoritative in-context knowledge was provided, which is analogous to the mechanism of retrieval-augmented generation.^28^

Several caveats temper these findings. The context-rich prompting result was achieved on purpose-built items with clean numerical inputs; real clinical data introduces measurement noise, mixed dentition complexities, and anomalous presentations that were not tested. Furthermore, context-rich prompting presupposes that the clinician or system designer already possesses the correct classification rules, creating a paradox whereby those who would benefit most from LLM assistance are least equipped to construct the prompts that would ensure accuracy. The use of consumer-facing web interfaces, whilst reflecting realistic clinical access modalities, precludes control over inference parameters and introduces reproducibility threats from undisclosed server-side updates.^29^ The absence of a non-thinking baseline condition means that the specific contribution of extended reasoning cannot be isolated. The binary scoring rubric does not capture error magnitude, and the synthetic data may not fully generalise to clinical practice.

## Conclusion

Frontier reasoning LLMs achieved high overall accuracy (94.0 to 99.0%) on orthodontic diagnostic computation but retained concealed, task-specific vulnerabilities detectable only through repeated-trial evaluation. Each model exhibited a distinct error profile, and these failures were concentrated at the normal-abnormal classification boundary. ChatGPT 5.4 Thinking showed deterministic cephalometric misclassification, Gemini 3 (Thinking) demonstrated confusion in within-grade IOTN-DHC sub-classification, and Claude Opus 4.6 (Extended Thinking) exhibited stochastic arithmetic variability. The observed tension between accuracy and consistency, whereby the most accurate model was the least consistent and the least accurate because its errors were the most deterministic, shows that single-trial evaluations are insufficient for characterising clinical risk. Context-rich prompting eliminated all errors on the study’s synthetic dataset, indicating that these failures reflected remediable parametric imprecision rather than fundamental reasoning incapacity. However, context-rich prompting should be regarded as a necessary but insufficient prerequisite for clinical deployment until prospectively validated against noisy, real-world patient records. Should clinical deployment ever be considered, standardised prompt templates incorporating authoritative classification criteria, together with multi-model verification and prior prospective validation, should be regarded as prerequisites.

## Data Availability

All data produced in the present work are contained in the manuscript.

## Author Contributions Statement

KWJ - Study design, test item construction, data collection, data analysis, manuscript drafting

JS - Study design, manuscript review, supervision

ELE - manuscript review and supervision

ALCY - manuscript review, supervision

KWCF - Study conception, study design, independent scoring, supervision, manuscript review and revision, corresponding author.

## References

1. Nordblom NF, Buttner M, Schwendicke F. Artificial Intelligence in Orthodontics: Critical Review. J Dent Res. 2024;103(6):577–84.

2. Gracea RS, Winderickx N, Vanheers M, Hendrickx J, Preda F, Shujaat S, et al. Artificial intelligence for orthodontic diagnosis and treatment planning: A scoping review. J Dent. 2025;152:105442.

3. Bedi S, Liu Y, Orr-Ewing L, Dash D, Koyejo S, Callahan A, et al. Testing and Evaluation of Health Care Applications of Large Language Models: A Systematic Review. JAMA. 2025;333(4):319–28.

4. Hatia A, Doldo T, Parrini S, Chisci E, Cipriani L, Montagna L, et al. Accuracy and Completeness of ChatGPT-Generated Information on Interceptive Orthodontics: A Multicenter Collaborative Study. J Clin Med. 2024;13(3).

5. Makrygiannakis MA, Giannakopoulos K, Kaklamanos EG. Evidence-based potential of generative artificial intelligence large language models in orthodontics: a comparative study of ChatGPT, Google Bard, and Microsoft Bing. Eur J Orthod. 2025;48(1).

6. Wafaie K, Basyouni ME, Bhattacharjee T, Prasad S, Daraqel B, Mohammed H. Diagnostic accuracy of generative large language artificial intelligence models for the assessment of dental crowding. BMC Oral Health. 2025;25(1):1558.

7. Banerjee S, Agarwal A, Singla S, editors. Llms will always hallucinate, and we need to live with this. Intelligent Systems Conference; 2025: Springer.

8. Shrestha S, Kim M, Ross K. Mathematical reasoning in large language models: Assessing logical and arithmetic errors across wide numerical ranges. arXiv preprint arXiv:250208680. 2025.

9. Boye J, Moell B. Large language models and mathematical reasoning failures. arXiv preprint arXiv:250211574. 2025.

10. Suh CH, Yi J, Shim WH, Heo H. Insufficient Transparency in Stochasticity Reporting in Large Language Model Studies for Medical Applications in Leading Medical Journals. Korean J Radiol. 2024;25(11):1029–31.

11. Davis J, Van Bulck L, Durieux BN, Lindvall C. The Temperature Feature of ChatGPT: Modifying Creativity for Clinical Research. JMIR Hum Factors. 2024;11:e53559.

12. Klishevich E, Denisov-Blanch Y, Obstbaum S, Ciobanu I, Kosinski M. Measuring determinism in large language models for software code review. arXiv preprint arXiv:250220747. 2025.

13. Snell C, Lee J, Xu K, Kumar A. Scaling llm test-time compute optimally can be more effective than scaling model parameters. arXiv preprint arXiv:240803314. 2024.

14. Jaech A, Kalai A, Lerer A, Richardson A, El-Kishky A, Low A, et al. Openai o1 system card. arXiv preprint arXiv:241216720. 2024.

15. Vrdoljak J, Boban Z, Males I, Skrabic R, Kumric M, Ottosen A, et al. Evaluating large language and large reasoning models as decision support tools in emergency internal medicine. Comput Biol Med. 2025;192(Pt B):110351.

16. Lewis P, Perez E, Piktus A, Petroni F, Karpukhin V, Goyal N, et al. Retrieval-augmented generation for knowledge-intensive nlp tasks. Advances in neural information processing systems. 2020;33:9459–74.

17. Wang L, Chen X, Deng X, Wen H, You M, Liu W, et al. Prompt engineering in consistency and reliability with the evidence-based guideline for LLMs. NPJ Digit Med. 2024;7(1):41.

18. Tay JRH, Chow DY, Lim YRI, Ng E. Enhancing patient-centered information on implant dentistry through prompt engineering: a comparison of four large language models. Front Oral Health. 2025;6:1566221.

19. Brook PH, Shaw WC. The development of an index of orthodontic treatment priority. Eur J Orthod. 1989;11(3):309–20.

20. Bolton WA. Disharmony in tooth size and its relation to the analysis and treatment of malocclusion. The Angle Orthodontist. 1958;28(3):113–30.

21. Bolton WA. The clinical application of a tooth-size analysis. American Journal of Orthodontics. 1962;48(7):504–29.

22. Kirschen RH, O’Higgins E A, Lee RT. The Royal London Space Planning: an integration of space analysis and treatment planning: Part I: Assessing the space required to meet treatment objectives. Am J Orthod Dentofacial Orthop. 2000;118(4):448–55.

23. Cooke MS, Wei SH. Cephalometric standards for the southern Chinese. Eur J Orthod. 1988;10(3):264–72.

24. Steiner CC. Cephalometrics for you and me. American journal of orthodontics. 1953;39(10):729–55.

25. Jacobson A. The “Wits” appraisal of jaw disharmony. Am J Orthod. 1975;67(2):125–38.

26. Liu M, Okuhara T, Dai Z, Huang W, Gu L, Okada H, et al. Evaluating the Effectiveness of advanced large language models in medical Knowledge: A Comparative study using Japanese national medical examination. Int J Med Inform. 2025;193:105673.

27. Huang L, Yu W, Ma W, Zhong W, Feng Z, Wang H, et al. A survey on hallucination in large language models: Principles, taxonomy, challenges, and open questions. ACM Transactions on Information Systems. 2025;43(2):1–55.

28. Nori H, King N, McKinney SM, Carignan D, Horvitz E. Capabilities of gpt-4 on medical challenge problems. arXiv preprint arXiv:230313375. 2023.

29. Chen L, Zaharia M, Zou J. How is ChatGPT’s behavior changing over time? Harvard Data Science Review. 2024;6(2).

